# On the timing of interventions to preserve hospital capacity: lessons to be learned from the Belgian SARS-CoV-2 pandemic

**DOI:** 10.1101/2020.12.18.20248450

**Authors:** Niel Hens, Christel Faes, Marius Gilbert

## Abstract

Using publicly available data on the number of new hospitalisations we use a newly developed phase portrait to monitor the epidemic allowing for assessing whether or not intervention measures are needed to keep hospital capacity under control. Using this phase portrait, we show that intervention measures were effective in mitigating a Summer resurgence but that too little too late was done to prevent a large autumn wave in Belgium.

## Introduction

Despite differences testing strategies as well as controversy with respect to (over)counting deaths, Belgium has been hit particularly hard by the coronavirus placing the country near the top in international rankings when looking at the official number of confirmed cases per 100,000 and the official number of deaths per million. On 6/12/2020, Belgium accounted for more than half a million confirmed cases and over 17,000 SARS-CoV-2 confirmed and suspected deaths. There are several factors explaining the vulnerability of Belgium to the SARS-CoV-2 pandemic including Belgium’s location at the centre of Europe and with Brussels being the capital of the European Union resulting in high international mobility as well as a high population density, high average household size and an older population structure that combined with a relatively high mixing behaviour increases transmission potential and the associated disease burden (1,2).

Belgium has known three surges of the coronavirus in 2020. The large number of hospitalizations of covid-19 patients has twice forced hospitals to postpone regular care of non-covid-19 patients. The first wave occurred between March 8 and June 1, accounting for a total of 58,641 confirmed cases with testing mostly focusing on severe illness, 17,132 hospitalizations and 9,377 deaths. The median age of hospitalized patients was 70 years (IQR 55-82). In the summer period, between July 1 and August 31, a local increase in confirmed cases was observed in the province of Antwerp and Brussels (24,056 confirmed cases in Belgium, of which 49.6% occurred in Antwerp and Brussels). It was mainly younger people who got infected in this period (median age 52 years, IQR 33-76), resulting in less severe infections and a smaller number of hospitalizations (1,220 hospitalizations), but it did put high pressure on general practitioners. A second large wave started on October 1, with 455,442 confirmed cases, 22,126 hospitalizations and 6,817 deaths on November 30. While confirmed cases are younger in this time period (median age 43, IQR 27-59), the age of hospitalisation is similar as in the first wave (median age 71, IQR 57-82). Changes in the testing strategy over time make comparisons of the number of confirmed cases difficult, but the number of hospitalizations is a more stable and important indicator of the severity of the outbreak and has a direct impact on the hospital capacity (3).

## Methodology

Using publicly available data from Sciensano on the number of new hospitalisations we define a phase portrait to monitor the epidemic allowing for assessing whether or not intervention measures are needed to keep hospital capacity under control (4). The diagram uses the 7-day average new hospitalizations and the daily ratio of the past 14-days new hospitalizations. For each combination, the total number of hospitalizations is projected for a 14-days horizon, from which the number of patients requiring intensive care is predicted based on the distribution of time spent in ICU (5, see appendix).

The hospital contingency plan in Belgium consists of 5 different phases while focusing on covid-19 related ICU care: Phase 0: 303 ICU-beds; Phase 1A: 528 ICU-beds; Phase 1B: 987 ICU-beds; Phase 2A: 1502 ICU-beds; Phase 2B: 2019 ICU-beds. Note that within this scheme the total number of patients (covid-19 and non-covid-19) in ICU moves from 2001 (Phase 0, 1A & 1B) to 2304 (Phase 2A) and 2821 (Phase 2B) and consequently yields a gradual decrease in non-covid-19 ICU capacity.

The cliquets’ diagram shows - from green to red - the severity of the outbreak in terms of hospital and future covid-related ICU load. The green region can be considered a “safe zone” in which the number of new hospitalisations is limited with a decrease (growth <1) or a limited increase (growth >1). This zone is associated with a limited number of covid-19 patients at ICU (<50 ICU beds in the next 14 days, a somewhat ad-hoc choice for the first part of phase 0). Next is the yellow region, a region of increased vigilance (second part of phase 0). The orange (phase 1A & 1B) and red (phase 2A & 2B) regions are “high impact” and “no-go” zones, in which non-covid-19 care decreases substantially and additional capacity for covid-19 needs to be provided for.

## Results

### Current situation

The current situation is presented in Figure 1 with starting point 1/10/2020. The situation in Belgium did worsen quickly and hospital networks moved from Phase 0 to Phase 1A (yellow to orange in the diagram), from Phase 1A to Phase 1B (dark orange) and eventually to Phase 2A (red). Intervention measures were implemented on October 19 and additional measures on October 24 and November 2, resulting in a slowing down of the growth of new hospitalizations followed by a decrease in new hospitalizations. Such a wave is described by a clockwise circular movement on the diagram. On November 16, primary schools and first grade secondary schools were fully reopened while second and third grade schools were partially reopened. About two weeks later, the growth in hospitalizations increased again, resulting in an upward movement in the diagram from the beginning of December.

**Figure 1:**
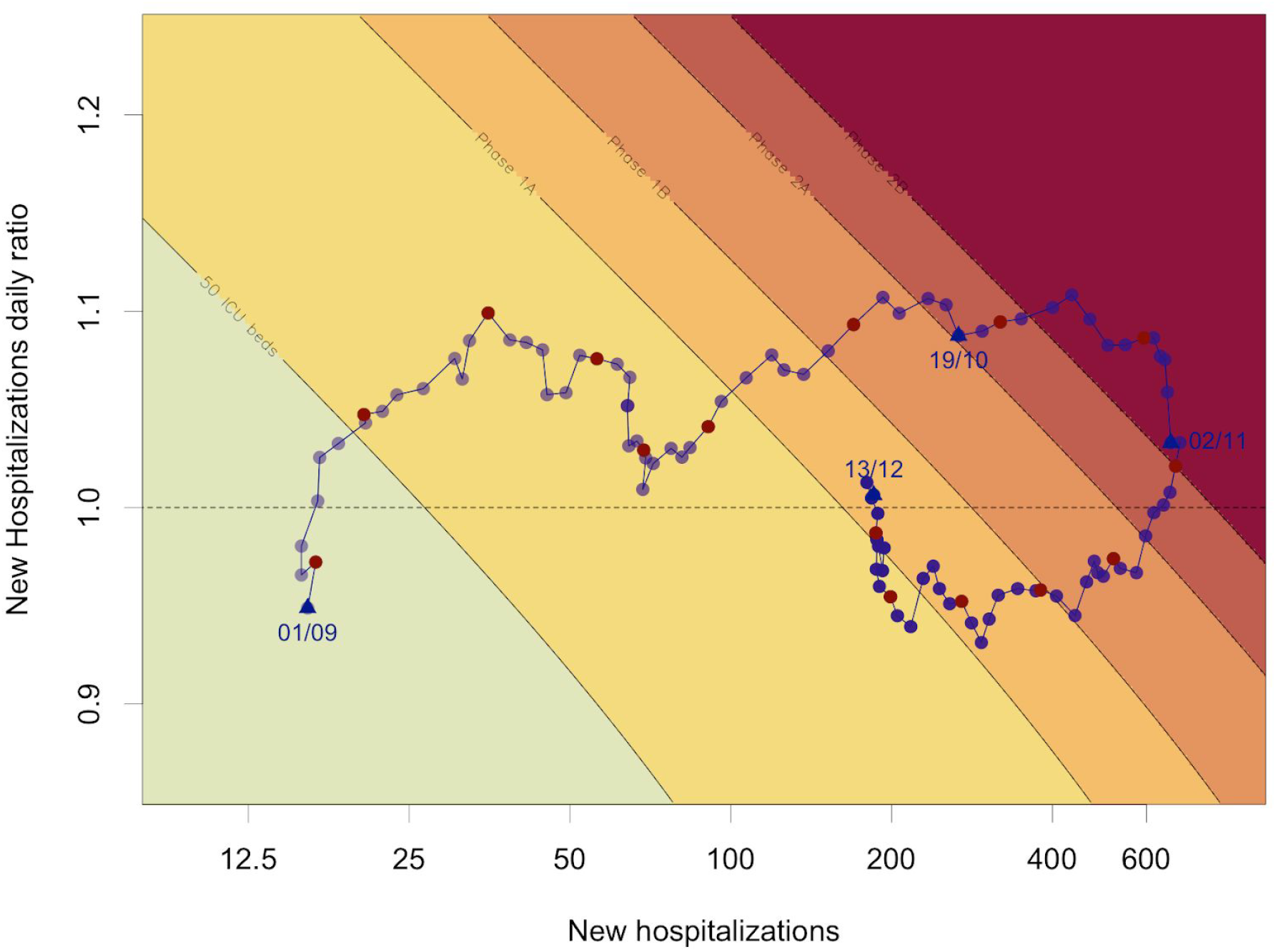
cliquets’ diagram of current situation from 01/10/2020 to 13/12/2020. Red dots correspond to Wednesdays.

### Learning from the past

A key question is whether the resurgence in hospitalisations could have been foreseen. Therefore we look at the phase portraits over three consecutive time periods throughout the pandemic (Figure 2). The first exit strategy was carefully designed (for a general overview of the main principles see 6) and consisted of 4 exit-phases (exit-phase 1a: May 5, exit-phase 1b: May 11, exit-phase 2: June 8, exit-phase 3: June 15, exit-phase 4: July 1). This exit strategy was largely successful in reducing the number of hospitalisations and keeping those numbers under control until July 23 (Figure 2 A,B). Thereafter the increase in the number of hospitalisations followed an asynchronous increase of the number of confirmed cases in the Provinces of Antwerp (starting July 12) and Brussels (starting July 27) which was successfully curbed in August following the instalment of additional control policies on July 27 at the national level and more strict measures in the province of Antwerp on July 28, translating into a loop in the phase portrait (Figure 2B). So, early and strong interventions when the trajectory entered the yellow region were successful in curbing the epidemic and bringing it back under control. In the course of September, an increase in the number of new hospitalisations was observed again following an increase in the number of confirmed cases at the end of August (Figure 2C). However, this time, no new policy measures were taken until October 9 when the trajectory had entered the orange zone, and these measures did not result in a decrease in the number of new hospitalisations after which the autumn wave depicted in Figure 1 followed. A considerable decrease in the daily ratio of the number of new hospitalisations was observed late September though there is no clear reason why this occurred.

**Figure 2:**
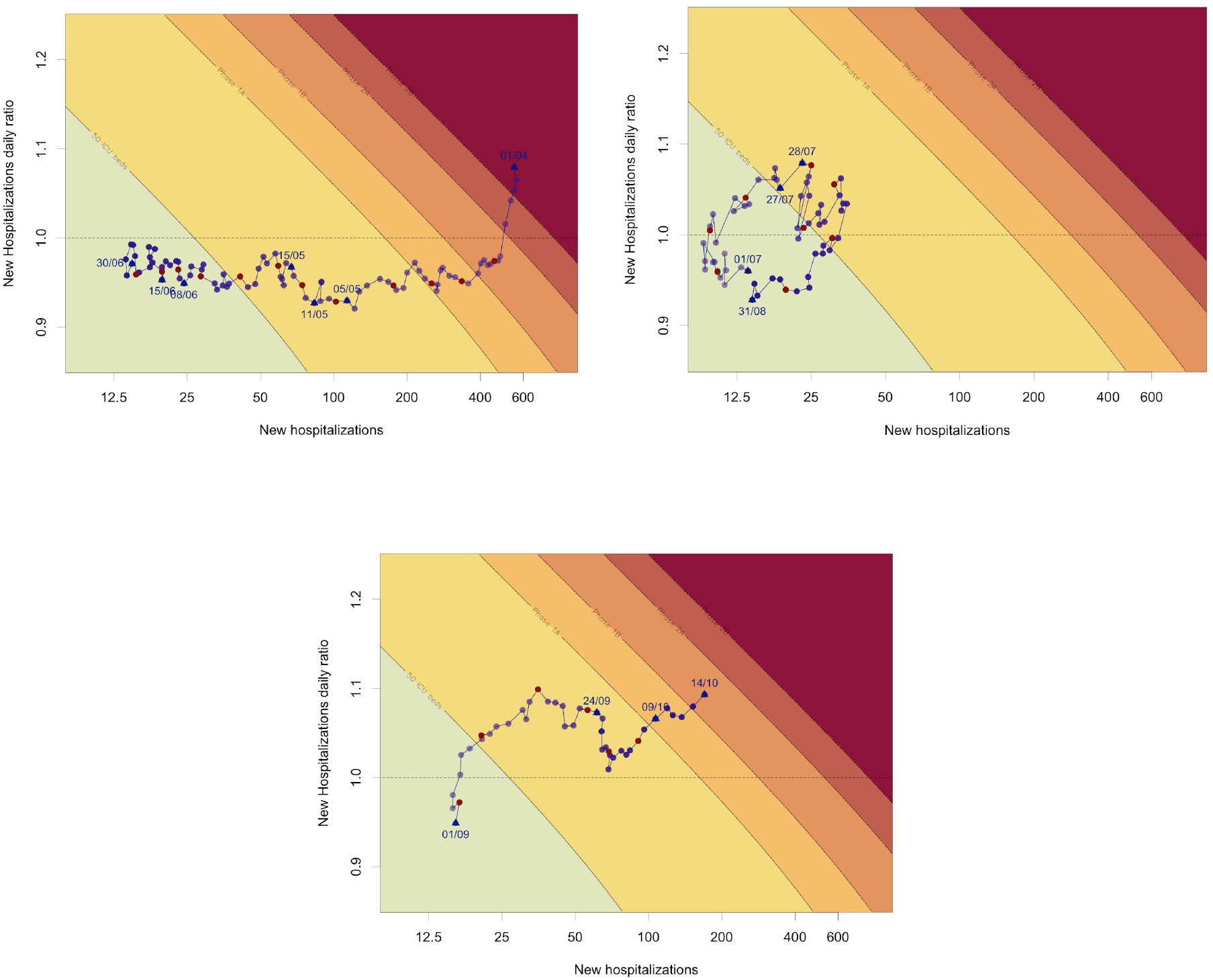
cliquets’ diagram for different time periods: (A) left upper panel: situation from April 1 to June 30, (B) right upper panel: situation from July 1 to August 31, (C) lower panel: situation from September 1 to October 14

## Discussion

We evaluated the SARS-CoV-2 pandemic in Belgium using a simple phase portrait depicting the number of new hospitalisations versus the daily ratio of new hospitalisations and predicting the number of covid-19 patients requiring intensive care.

Dividing the pandemic in different time periods and using the cliquets’ diagram clearly shows that the intervention measures in August, i.e. in the yellow area, were successful in gaining control over the pandemic. In September-October, however, there was a substantial increase in the number of new covid-19 hospitalisations whereas new non-pharmaceutical interventions only started when entering in the orange “high impact” area. Moreover, these interventions appeared to be insufficiently strong to curb the epidemic. It is important to note that the inaction in September-October coincided with a transition from a temporary federal government to the installment of a definitive one and a high level of scepticisms toward the reality of the restart of the epidemic by several experts in the social and conventional media.

The cliquets’ diagram is merely a visualisation of the epidemiological situation in hospitals. But it allows for simultaneously visualizing where we are in terms of speed (new hospitalizations per day) and acceleration (daily ratio) of the epidemic with a forward thinking toward the 14-days horizons covid-19 ICU occupancy. The historical situation shows that interventions taken early in the yellow region were successful in keeping the hospital capacity under control. Note that Belgium, relative to other countries, has a relatively large ICU capacity which likely leads to overconfidence in policy control whereas early intervention is key given that with low numbers mitigation strategies are much more effective.

There are several limitations related to the proposed cliquets’ diagram. First, we relied on the daily number of new covid-19 hospitalisations which are available for Belgium through a daily hospital surge survey developed and implemented by the national public health organization Sciensano and for which hospitals provided timely input (4). This may not be available for other countries. Second, using new hospitalisations yields a more stable, but somewhat late indicator. Combining the daily ratio based on, e.g., confirmed cases, gives a lead time, which we estimated to be 7-10 days, (results not shown). We believe delays and underreporting in the number of confirmed cases doesn’t have a large impact given that changing case definitions and test saturation are only likely to occur when already in a high-impact or no-go zone. Using test positivity rates could provide a useful addition to the number of confirmed cases. Further research includes defining a phase portrait based on confirmed cases though the connection to the hospital contingency phases is less straightforward because of the age-specificity of hospitalisation rates.

## Data Availability

The data used in this manuscript has been made available by Sciensano.

https://epistat.wiv-isp.be/covid/

## Acknowledgments

We thank Sciensano for making their hospital data publicly available. NH and CF acknowledge support from the European Union’s Horizon 2020 research and innovation programme - project EpiPose (Grant agreement number 101003688) and NH acknowledges support from the European Research Council (ERC) under the European Union’s Horizon 2020 research and innovation programme (grant agreement 682540 - TransMID). We are grateful for comments received by Mathias Dewatripont, Geert Molenberghs, Philippe Beutels, Emmanuel André, Herman Goossens and Pierre Van Damme.

## Appendix

Publicly available data from Sciensano are used on the number of new hospitalisations, total number of hospital beds and total number of intensive care (ICU) beds, which are reported on a daily basis (www.sciensano.be).

Data are presented as the pair of new hospitalizations and growth rate of new hospitalizations. The new hospitalizations are averages of the past 14-day number of new hospitalizations. The growth rate is obtained from modeling the past 14-day number of new hospitalizations *H* (*t*) as

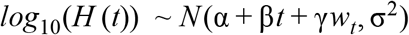

with *w*_*t*_ an indicator for weekends and holidays. The growth rate is then calculated as

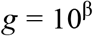

Colours of the phase diagram are based on the predicted ICU capacity. For an assumed number of new hospitalizations *H* (*t*_0_) on day *t*_0_and and assumed (constant) growth rate *g*, we calculate the number of new hospitalizations in the next *t* days as

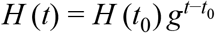

Based on the predicted number of new hospitalisations, we derive the total number of patients in intensive care unit (ICU), based on the proportion of hospitalised patients that need intensive care and the length of stay in ICU. The number of patients in ICU on day *t* is

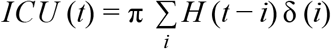

where π is the fraction of hospitalized patients going to ICU and δ (*i*) is the probability that a patient stays *i* days in ICU.

The distribution of length of stay in ICU is derived from a multicenter registry in Belgium that collects information on hospital admission related to COVID-19 infection, for the period March to August 2020 (Faes et al. 2020, Van Goethem et al 2020). Patients staying longer than 60 days in ICU are assumed to leave ICU on day 60. The fraction π of hospitalized patients that need intensive care is, based on the survey, 0.152. As this is a survey, and does not contain all hospitalizations, this is corrected by the fraction of daily ICU beds reported in the hospital survey versus the daily reported national ICU load (1/2.15) and the fraction of hospital beds reported in the survey and reported nationally (1/1.33). As a result, we estimate π as 0.245.

As cutpoints, we use the estimated ICU load on day *t*_0_ + 14 (which resembles the covid-19 ICU load if the behavior stays the same in 2 weeks). Cutpoints are chosen in correspondence to the hospital contingency plan in Belgium consisting of 5 different phases while focusing on covid-19 related ICU care: Phase 0: 303 ICU-beds; Phase 1A: 528 ICU-beds; Phase 1B: 987 ICU-beds; Phase 2A: 1502 ICU-beds; Phase 2B: 2019 ICU-beds. Note that within this scheme the total number of patients (covid-19 and non-covid-19) in ICU moves from 2001 (Phase 0, 1A & 1B) to 2304 (Phase 2A) and 2821 (Phase 2B) and thus non-covid-19 care gradually decreases with an increase in phases.

The code is available at https://github.com/NielHens/Cliquets.

